# Protocol: Waiting time and ways of accessing specialized health services in public hospitals in Ecuador

**DOI:** 10.1101/2024.11.25.24317843

**Authors:** Marcelo Armijos Briones, Sammy Figueroa Intriago, Antonio Lanata Flores, Pablo Benitez Sellán, Oscar Marcillo Toala, Patricia Estefanía Ayala Aguirre

**Author notes:** Conceptualization; Formal Analysis; Methodology; Writing – Original Draft Preparation (Marcelo Armijos Briones). Validation; Visualization; Writing – Review & Editing (Sammy Figueroa Intriago, Antonio Lanata Flores, Pablo Benitez Sellán, Oscar Marcillo Toala, Patricia Estefanía Ayala Aguirre). Funding: This research project is 100% funded by the Research Center of the University of Specialties Espíritu Santo. This institution is the same as that of all the authors of this protocol. Competing interests: the authors declare that they have no conflict of interest during the development of this research protocol. Data availability: the data of this protocol are registered and publicly accessible at Protocols.io https://dx.doi.org/10.17504/protocols.io.261ge5z7wg47/v1 with Protocol Integer ID 109306. The data collected will be made available upon request after publication, following journal policies. After that, that is, when the protocol is accepted for publication, the data collection process will begin. This is possible because the approval of the ethics committee has already been obtained.

## Abstract

This study aims to determine the waiting time and the forms of access to specialized health services in public hospitals in Ecuador. A representative sample of 32 hospitals under the Ministry of Public Health was considered, with 26 selected by accessibility convenience. Data will be collected using a structured questionnaire. Patients will be asked about the number of days they waited for their medical appointments and the method used to schedule their appointments. The study distinguishes between standardized access, based on Ecuador’s formal referral and counter-referral system, and non-standardized access methods, such as personal connections or hospital staff involvement. The data of this protocol are registered and publicly accessible at: https://dx.doi.org/10.17504/protocols.io.261ge5z7wg47/v1

We expect to identify a correlation between waiting times and the type of access to specialized medical services, with non-standardized access potentially leading to shorter waiting times. This research may highlight disparities in the system and suggest areas for improvement in equity and efficiency within the healthcare referral process. To do so, a structured survey will be used. Since a construct is not needed to determine the waiting time or the forms of access, it was not necessary to validate the instrument. However, we did validate the understanding of the questions and the response options in several places in the country. According to the results According to the results of the validation of the instrument, pollsters will be instructed to inform users about the meaning of the question on ethnic identification, which was difficult to understand in the country’s coastal areas.

## Background

Ecuador’s health system is characterized by a mix of public and private entities, with the Ministry of Public Health (MSP) playing a central role in its operation and evaluation.

Even though the Constitution recognizes health as a human right in the whole Comprehensive Public Health Network, there are challenges that need to be addressed such as: extensive/long working hours (or shifts), availability and quality of medicines and a perceived disconnect between governance and medical service (1,2). Despite constitutional guarantees for healthcare access, the dominance of the biomedical model and top-down policy enforcement have led to issues in addressing health epidemics in rural areas (3). Faith-based healthcare providers (FBHPs) have historically contributed in parallel with the system, but their current role in the national health care requires further understanding (4). In summary, Ecuador’s health system is a complex interplay of the public and private sectors, with constitutional commitments to health as a human right and ongoing efforts to improve public health infrastructure. However, the system faces significant challenges, including resource limitations, healthcare provider burnout, and the need for better integration of various health services and innovative systems. Addressing these issues is crucial for the system’s reliability and health outcomes in the Ecuadorian population (5). For example, to improve system reliability, Ecuador created the national referral and counter-referral subsystem, which, among other things, aims to make specialized care more efficient, leaving the first level of care as the gateway to the national health system and regulating access to specialized health services for those who need it.

The referral and counter-referral system in Ecuador, as described in the context provided, is part of a healthcare framework that was reformed following the 2008 Constitution, which guaranteed access to healthcare for all citizens (3). This system involves the process of referring patients from lower-capacity institutions to higher-complexity institutions for specialized care and diagnoses and then counter-referring them back to the original institution with specific diagnoses and treatment plans (6). In summary, the referral and counter-referral system in Ecuador is a critical component of the healthcare process designed to facilitate the movement of patients through different levels of care. However, the effectiveness of the system is hindered by the prevailing healthcare model and administrative practices that do not adequately support continuous and community-based care (3,7).

With this protocol, we intend to test the hypothesis that the way of accessing health services intervenes with the waiting time in public hospitals. There is local evidence that this phenomenon is happening, but the national dynamics have not been studied systematically. Inquiry into waiting times and forms of access to specialized medical services in public hospitals in Ecuador is a multifaceted issue. This research aims to discern the relationship between the mode of access, whether through formal channels or informal networks—and the duration patients wait for specialized consultations. The context provided by Briones et al. (2024) is particularly relevant, as it directly investigates waiting times in Ecuadorian public hospitals and identifies informal access as a potential factor in scheduling delays (8). Contradictory findings emerge when considering the broader literature on waiting times for health care services. While some studies suggest that socioeconomic status does not significantly affect waiting times for certain services (9), others indicate that lower socioeconomic status is associated with longer waits (10,11). In summary, the relationship between access forms and waiting times for specialized medical services in Ecuadorian public hospitals is complex and is influenced by various factors, including informal access networks, socioeconomic status, and external events. Evidence suggests that while formal processes may lead to longer waiting times, informal networks can expedite access, albeit potentially exacerbating inequities (8). To fully understand the dynamics at play, it is crucial to consider the broader context of healthcare access and waiting times, as illuminated by the referenced international studies.

Understanding the time that users of public hospitals in Ecuador wait for specialized medical care is crucial for improving the country’s healthcare system. This information can help policymakers and healthcare providers identify areas that require improvement and allocate resources more effectively. Previous studies have shown that patients in Ecuador often face long waiting times for medical appointments, with some patients waiting months to see a specialist. However, there is limited research on the specific time that users of public hospitals in Ecuador wait for specialized medical care. There is a lack of research on the time that users of public hospitals in Ecuador wait for specialized medical care and how they access these services. This study aims to fill this knowledge gap by providing detailed information on waiting times and referral systems in Ecuadorian public hospitals.

This study is essential for understanding the challenges faced by the Ecuadorian healthcare system and identifying ways to improve the delivery of specialized medical care. This study provides valuable insights for policymakers and healthcare providers by examining waiting times and referral systems in public hospitals. What is the average waiting time for specialized medical care in public hospitals in Ecuador and how are patients accessing these services?

This study aimed to determine the average waiting time for specialized medical care in public hospitals in Ecuador and to examine the referral and counter-referral systems used by patients to access these services. It is hypothesized that patients in public hospitals in Ecuador face long waiting times for specialized medical care, and the referral and counter-referral systems do not efficiently manage patient appointments.

## Method

The research design for this study is a cross-sectional survey design. The research method used in this study is a structured questionnaire survey. The study participants will be patients who have undergone specialized medical consultations in the 26 public hospitals in Ecuador. We will wait to begin conducting surveys outside public hospitals until the article with the protocol is approved. We hope that this will be in January or February 2025. The approval obtained by the ethics committee lasts one year, until August 2025. If necessary, it is possible to request an extension. Before filling out the surveys, participants must agree to participate after the interviewer reads them the informed consent that is present in digital form in the Google form. Participation by minors or persons who are not in suitable intellectual capacities will not be accepted, due to this, the consent of legal guardians is not necessary.

Inclusion Criteria: Inclusion criteria for this study will be patients who have undergone specialized medical consultations in the 26 public hospitals in Ecuador.

Exclusion Criteria: Exclusion criteria for this study will be patients who are unable to provide informed consent, patients who are unable to understand the questionnaire, and patients who have undergone specialized medical consultations in private hospitals.

For the sample calculation in each hospital, the Epi Info application was used with the following parameters: confidence level 95%, margin of error 5%, event prevalence 82% because, according to the Briones 2024 study, this number of appointments in public hospitals in the province of Manabí in Ecuador are by standardized scheduling. The number of visits to the Outpatient Clinic areas, the area where specialized medical care is provided in the country’s public hospitals, was taken from the data of the Ecuadorian Ministry of Public Health on its website. The morbidity care data for the month of August 2023 was taken as a reference whenever data from that year existed; when not, they were taken from the last year available as shown in Table 1.

**Table 1.** Stratification of proportional sampling of users who attend medical consultations of specialties in public hospitals in Ecuador

|  |  |  |  |  |
| --- | --- | --- | --- | --- |
|  | Hospital | Number of morbidity care visits in August | Year | Sample |
| 1 | Alfredo Noboa Montenegro General Hospital | 2289 | 2022 | 200 |
| 2 | Ambato General Hospital | 11858 | 2023 | 215 |
| 3 | Dr. Verdi Cevallos Balda General Hospital | 7212 | 2023 | 212 |
| 4 | Enrique Garces General Hospital | 9748 | 2023 | 214 |
| 5 | Esmeraldas South General Hospital – “Delfina Torres de Concha” | 3454 | 2023 | 206 |
| 6 | Francisco de Orellana General Hospital | 3450 | 2022 | 206 |
| 7 | Guasmo South General Hospital | 1908 | 2016 | 196 |
| 8 | Gustavo Dominguez General Hospital | 6423 | 2023 | 212 |
| 9 | Homero Castanier Crespo General Hospital | 1581 | 2016 | 192 |
| 10 | Isidro Ayora General Hospital | 508 | 2021 | 153 |
| 11 | José María Velasco Ibarra General Hospital | 6169 | 2023 | 211 |
| 12 | Julius Doepfner General Hospital | 1772 | 2023 | 195 |
| 13 | Latacunga General Hospital | 10468 | 2023 | 214 |
| 14 | León Becerra General Hospital | 2662 | 2022 | 202 |
| 15 | Liborio Panchana Sotomayor General Hospital | 3527 | 2023 | 206 |
| 16 | Luis Gabriel Davila General Hospital | 3263 | 2023 | 205 |
| 17 | Hospital General Macas General Hospital | 2566 | 2022 | 202 |
| 18 | Marco Vinicio Iza General Hospital | 4969 | 2023 | 210 |
| 19 | General Martin Icaza General Hospital | 3152 | 2022 | 205 |
| 20 | Pablo Arturo Suarez General Hospital | 14388 | 2023 | 216 |
| 21 | Hospital General Puyo General Hospital | 4319 | 2023 | 208 |
| 22 | General Rodriguez Zambrano General Hospital | 1876 | 2023 | 196 |
| 23 | Vicente De Paul General Hospital | 5680 | 2023 | 211 |
| 24 | General Teofilo Davila General Hospital | 5577 | 2023 | 211 |
| 25 | Vicente Corral Moscoso General Hospital | 2187 | 2020 | 199 |
| 26 | General Teaching Provincial Hospital of Riobamba | 6946 | 2023 | 212 |
| Total |  | 127952 |  | 5309 |

### Data Collection

The data collection instrument was a semi-structured survey containing a total of 30 questions with an average duration of 8 minutes to be answered. The questions were divided into several sections, the first containing social, demographic and economic information, the second containing ethnic and cultural information and the third containing information on the method of access and waiting time for specialized medical care. For the first and second sections, questions taken from the Health and Nutrition survey forms carried out periodically by the Institute of Statistics and Census of Ecuador (INEC) were used. Because of this, these questions did not need to be validated since they are essentially descriptive questions such as age, sex, place of residence, among others, as shown below in the table 2:

**Table 2.** Questions and response options for the sections: social, demographic and economic information and ethnic and cultural information

|  |  |
| --- | --- |
| Questions from the first section in English | Answer options |
| Age | Number of years of life completed up to the date of the survey |
| Sex | Female or male |
| Level of formal education achieved | None, primary, secondary, third level or fourth level |
| What is the average income of your household (insert the approximate figure of the income of all the people who live in your house, an exact value is not necessary) | Amount of money in dollars that the family receives per month |
| How many people live in your house (insert the number of people who live in the respondent's home) | Number of people living permanently in their home |
| Area of residence | Urban or rural |
| Do you live in the same province as the hospital? | Yes or no |
| If not, why were you not treated in your province? | The specialist I need is not available; this hospital is closer to my residence; a doctor made an appointment for me at this hospital; other reason |
| Province of residence | Any of the 23 continental provinces of Ecuador |
| From your residence to the hospital, how long did it take you to get to the hospital? (insert the data in minutes) | The time in minutes it took to get to the hospital |
| In total, how much did you spend on transportation to get to the hospital? (insert the dollar value of the round-trip expense per person) | The amount in dollars spent to get to the hospital |
| Ethnic self-identification (ask the respondent what their ethnicity is) | Mestizo, white, indigenous, black, Afro-Ecuadorian, mulatto, montuvio, other |
| What is your native language (explain that this is your original language, the one you use as your first language to communicate at home) | Kichwua; Waorani; Shuar; Achuar; Cha'pala; Awapit; Spanish; other |
| Write the other language | The name of the other language mentioned by the patient is written verbatim |
| Did anyone at the hospital you spoke to know your language? (refers to someone who told you something about your medical appointment) | Yes or no |
| Do you think your ethnicity is an obstacle when scheduling an appointment? (to understand if during the scheduling process you had difficulties due to your language or ethnicity) | Yes; no; maybe |
| Do you think your native language is an obstacle when receiving medical care? (to understand if in previous appointments you had difficulty explaining your situation to the specialist) | Yes; no; maybe |

For the third section, questions were asked about the forms of access and the waiting time for specialized health services. In this section, we began by asking the reason for attending the hospital, because, in the area of specialty consultation, specialized procedures such as ultrasounds, x-rays, among others, are also considered and we were interested in measuring the waiting time for these procedures as well. The questions in this section are similar to those applied in the INEC Questionnaire in its national surveys, however, when modified to meet the objective of this research, they went through a process of validation of the clarity and consistency of the questions asked by the research team and related researchers. The process was carried out with a pilot study in three hospitals in the country, one in each geographic region (coast, mountains and Amazon). A total of 322 people were surveyed in hospitals in the 3 geographic regions of the country, who answered two questions after answering the questionnaire: Was there any question that you did not understand? and Did any question seem confusing or ambiguous? With yes or no response options. When the answer was Yes, the question why was asked. The answers were tabulated and analyzed with Pearson’s Chi Square test and Z Test for comparison of proportions. Table 3 describes the social, demographic and cultural characteristics of the participants in the validation.

**Table 3.** Characteristics of the population participating in the validation of the consistency of the questions in the questionnaire (n 322)

|  | N | % |
| --- | --- | --- |
| **Total** | 322 | 100,0 |
| **Gender** |  |  |
| Female | 213 | 66,1 |
| Male | 109 | 33,9 |
| **Age group** |  |  |
| 18 to 25 years | 40 | 12,4 |
| 26 to 40 years | 124 | 38,5 |
| 41 to 55 years | 95 | 29,5 |
| 56 to 70 years | 53 | 16,5 |
| 71 years and older | 10 | 3,1 |
| **Income quintile** |  |  |
| Quintile 1 | 105 | 32,6 |
| Quintile 2 | 156 | 48,4 |
| Quintile 3 | 45 | 14 |
| Quintile 4 | 11 | 3,4 |
| Quintile 5 | 5 | 1,6 |
| **Education** |  |  |
| None | 14 | 4,3 |
| Primary | 99 | 30,7 |
| Secondary | 167 | 51,9 |
| Third level | 40 | 12,4 |
| Fourth level | 2 | 0,6 |
| **Area of residence** |  |  |
| Urban | 251 | 78,0 |
| Rural | 71 | 22,0 |
| **Ethnicity** |  |  |
| Mestizo | 299 | 92,9 |
| Indigenous | 12 | 3,7 |
| Afro-ecuadorian | 2 | 0,6 |
| Montuvio | 9 | 2,8 |
| **Language** |  |  |
| Spanish | 311 | 96,6 |
| Kichwua | 6 | 1,9 |
| Shuar | 5 | 1,6 |

| **Geographic region** |  |  |
| --- | --- | --- |
| Sierra | 77 | 23,9 |
| Coast East | 110 | 34,2 |
| Oriente | 135 | 41,9 |

A balance was sought among participants due to the cultural disparity between regions in Ecuador. The results of the responses with the statistical tests mentioned above are shown in Table 4 and 5.

**Table 4.** Responses to the question “Were there any questions you did not understand?” from the people who participated in the validation of the consistency and understanding of the questions that made up the survey applied

|  | Were there any questions you did not understand? (n 324) |  |  |  |  |  | pvalor |
| --- | --- | --- | --- | --- | --- | --- | --- |
|  | No |  | Yes |  | Total |  |  |
|  | N | % | N | % | N | % |  |
| **Gender** |  |  |  |  |  |  | 0,131<sup>†</sup> |
| Female | 188<sub>a</sub> | 88,3 | 25<sub>a</sub> | 11,7 | 213 | 100 |  |
| Male | 102<sub>a</sub> | 93,6 | 7<sub>a</sub> | 6,4 | 109 | 100 |  |
| **Age group** |  |  |  |  |  |  | 0,344<sup>†</sup> |
| 18 to 25 years | 36<sub>a</sub> | 90 | 4<sub>a</sub> | 10 | 40 | 100 |  |
| 26 to 40 years | 107<sub>a</sub> | 86,3 | 17<sub>a</sub> | 13,7 | 124 | 100 |  |
| 41 to 55 years | 89<sub>a</sub> | 93,7 | 6<sub>a</sub> | 6,3 | 95 | 100 |  |
| 56 to 70 years | 48<sub>a</sub> | 90,6 | 5<sub>a</sub> | 9,4 | 53 | 100 |  |
| 71 years and older | 10<sub>a</sub> | 100 | 0<sub>a</sub> | 0 | 10 | 100 |  |
| **Income quintile** |  |  |  |  |  |  |  |
| Quintile 1 | 91<sub>a</sub> | 86,7 | 14<sub>a</sub> | 13,3 | 105 | 100 | 0,572<sup>‡</sup> |
| Quintile 2 | 143<sub>a</sub> | 91,7 | 13<sub>a</sub> | 8,3 | 156 | 100 |  |
| Quintile 3 | 42<sub>a</sub> | 93,3 | 3<sub>a</sub> | 6,7 | 45 | 100 |  |
| Quintile 4 | 10<sub>a</sub> | 90,9 | 1<sub>a</sub> | 9,1 | 11 | 100 |  |
| Quintile 5 | 4<sub>a</sub> | 80 | 1<sub>a</sub> | 20 | 5 | 100 |  |
| **Education** |  |  |  |  |  |  |  |
| None | 13<sub>a</sub> | 92,9 | 1<sub>a</sub> | 7,1 | 14 | 100 | 0,472<sup>‡</sup> |
| Primary | 93<sub>a</sub> | 93,9 | 6<sub>a</sub> | 6,1 | 99 | 100 |  |
| Secondary | 148<sub>a</sub> | 88,6 | 19<sub>a</sub> | 11,4 | 167 | 100 |  |
| Third level | 34<sub>a</sub> | 85 | 6<sub>a</sub> | 15 | 40 | 100 |  |
| Fourth level | 2<sub>a</sub> | 100 | 0<sub>a</sub> | 0 | 2 | 100 |  |
| **Area of residence** |  |  |  |  |  |  |  |
| Urban | 229<sub>a</sub> | 91,2 | 22<sub>a</sub> | 8,8 | 251 | 100 | 0,185<sup>†</sup> |
| Rural | 61<sub>a</sub> | 85,9 | 10<sub>a</sub> | 14,1 | 71 | 100 |  |
| **Ethnicity** |  |  |  |  |  |  |  |
| Mestizo | 267<sub>a</sub> | 89,3 | 32<sub>a</sub> | 10,7 | 299 | 100 | 0,434<sup>‡</sup> |
| Indigenous | 12<sub>a</sub> | 100 | 0<sub>a</sub> | 0 | 12 | 100 |  |
| Afro-ecuadorian | 2<sub>a</sub> | 100 | 0<sub>a</sub> | 0 | 2 | 100 |  |
| Montuvio | 9<sub>a</sub> | 100 | 0<sub>a</sub> | 0 | 9 | 100 |  |
| **Language** |  |  |  |  |  |  |  |
| Spanish | 279<sub>a</sub> | 89,7 | 32<sub>a</sub> | 10,3 | 311 | 100 | 0,533<sup>‡</sup> |
| Kichwua | 6<sub>a</sub> | 100 | 0<sub>a</sub> | 0 | 6 | 100 |  |
| Shuar | 5<sub>a</sub> | 100 | 0<sub>a</sub> | 0 | 5 | 100 |  |
| **Geographic region** |  |  |  |  |  |  |  |
| Sierra | 74<sub>a</sub> | 96,1 | 3<sub>b</sub> | 3,9 | 77 | 100 | 0,000<sup>†</sup> |
| Coast East | 85<sub>a</sub> | 77,3 | 25<sub>b</sub> | 22,7 | 110 | 100 |  |
| Oriente | 131<sub>a</sub> | 97 | 4<sub>b</sub> | 3 | 135 | 100 |  |

Each letter in the subscript denotes a subset of the Were there any questions you didn't understand? categories whose column proportions do not differ significantly from each other at the .05 level.
<sup>†</sup> Refers to the p-value of Pearso's chi-square test
<sup>‡</sup> Refers to the p-value of Fisher's Exact Test

**Table 5.** Responses to the question “Were any questions confusing or ambiguous?” from the people who participated in the validation of the consistency and understanding of the questions that made up the survey applied

|  | Were any questions confusing or ambiguous? (n 324) |  |  |  |  |  | Pvalor |
| --- | --- | --- | --- | --- | --- | --- | --- |
|  | No |  | Yes |  | Total |  |  |
|  | N | % | N | % | N | % |  |
| **Gender** |  |  |  |  |  |  | 1,000<sup>‡</sup> |
| Female | 208<sub>a</sub> | 97,7 | 5<sub>a</sub> | 2,35 | 213 | 100,00 |  |
| Male | 106<sub>a</sub> | 97,2 | 3<sub>a</sub> | 2,75 | 109 | 100,00 |  |
| **Age group** |  |  |  |  |  |  | 0,689<sup>‡</sup> |
| 18 to 25 years | 39<sub>a</sub> | 97,5 | 1<sub>a</sub> | 2,5 | 40 | 100,0 |  |
| 26 to 40 years | 119<sub>a</sub> | 96,0 | 5<sub>a</sub> | 4,0 | 124 | 100,0 |  |
| 41 to 55 years | 94<sub>a</sub> | 98,9 | 1<sub>a</sub> | 1,1 | 95 | 100,0 |  |
| 56 to 70 years | 52<sub>a</sub> | 98,1 | 1<sub>a</sub> | 1,9 | 53 | 100,0 |  |
| 71 years and older | 10<sub>a</sub> | 100,0 | 0<sub>a</sub> | 0,0 | 10 | 100,0 |  |
| **Income quintile** |  |  |  |  |  |  | 0,408<sup>‡</sup> |
| Quintile 1 | 102<sub>a</sub> | 97,1 | 3<sub>a</sub> | 2,9 | 105 | 100,0 |  |
| Quintile 2 | 152<sub>a</sub> | 97,4 | 4<sub>a</sub> | 2,6 | 156 | 100,0 |  |
| Quintile 3 | 45<sub>a</sub> | 100 | 0<sub>a</sub> | 0 | 45 | 100,0 |  |
| Quintile 4 | 10<sub>a</sub> | 90,9 | 1<sub>a</sub> | 9,1 | 11 | 100,0 |  |
| Quintile 5 | 5<sub>a</sub> | 100 | 0<sub>a</sub> | 0 | 5 | 100,0 |  |
| **Education** |  |  |  |  |  |  | 0,789<sup>‡</sup> |
| None | 14<sub>a</sub> | 100,0 | 0<sub>a</sub> | - | 14 | 100,0 |  |
| Primary | 97<sub>a</sub> | 98,0 | 2<sub>a</sub> | 2,0 | 99 | 100,0 |  |
| Secondary | 161<sub>a</sub> | 96,4 | 6<sub>a</sub> | 3,6 | 167 | 100,0 |  |
| Third level | 40<sub>a</sub> | 100,0 | 0<sub>a</sub> | - | 40 | 100,0 |  |
| Fourth level | 2<sub>a</sub> | 100,0 | 0<sub>a</sub> | - | 2 | 100,0 |  |
| **Area of residence** |  |  |  |  |  |  | 0,838<sup>†</sup> |
| Urban | 245<sub>a</sub> | 97,6 | 6<sub>a</sub> | 2,4 | 251 | 100,0 |  |
| Rural | 69<sub>a</sub> | 97,2 | 2<sub>a</sub> | 2,8 | 71 | 100,0 |  |
| **Ethnicity** |  |  |  |  |  |  | 1,000<sup>‡</sup> |
| Mestizo | 291<sub>a</sub> | 97,3 | 8<sub>a</sub> | 2,7 | 299 | 100,0 |  |
| Indigenous | 12<sub>a</sub> | 100,0 | 0<sub>a</sub> | 0,0 | 12 | 100,0 |  |
| Afro-ecuadorian | 2<sub>a</sub> | 100,0 | 0<sub>a</sub> | 0,0 | 2 | 100,0 |  |
| Montuvio | 9<sub>a</sub> | 100,0 | 0<sub>a</sub> | 0,0 | 9 | 100,0 |  |
| **Language** |  |  |  |  |  |  | 1,000<sup>‡</sup> |
| Spanish | 303<sub>a</sub> | 97,4 | 8<sub>a</sub> | 2,6 | 311 | 100,0 |  |
| Kichwua | 6<sub>a</sub> | 100,0 | 0<sub>a</sub> | 0,0 | 6 | 100,0 |  |
| Shuar | 5<sub>a</sub> | 100,0 | 0<sub>a</sub> | 0,0 | 5 | 100,0 |  |
| **Geographic region** |  |  |  |  |  |  | 0,664<sup>‡</sup> |
| Sierra | 74<sub>a</sub> | 96,1 | 3<sub>a</sub> | 3,9 | 77 | 100,0 |  |
| Coast East | 108<sub>a</sub> | 98,2 | 2<sub>a</sub> | 1,8 | 110 | 100,0 |  |
| Oriente | 132<sub>a</sub> | 97,8 | 3<sub>a</sub> | 2,2 | 135 | 100,0 |  |

Each letter in the subscript denotes a subset of the Were there any questions you didn't understand? categories whose column proportions do not differ significantly from each other at the .05 level.
<sup>†</sup> Refers to the p-value of Pearso's chi-square test
<sup>‡</sup> Refers to the p-value of Fisher's Exact Test

This type of validation was chosen because the purpose of this questionnaire is to measure the waiting time in days and to find out whether or not people use the referral and counter-referral system stipulated in the Ecuadorian Health Model, knowing the origin of the medical appointment. Both measurement scales were considered psychometric, that is, they can be measured objectively by asking the population directly (12). Because of this, it is not necessary to develop a series of questions to determine a construct. The latter is used when the measurement scales are psychophysical, such as when it is required to measure the level of satisfaction, the quality of medical care, among others.

In Table 4, it is possible to observe that in the coastal region or Ecuadorian littoral, the people who participated in the validation had some difficulties when understanding some of the questions. When they were asked about what the difficulty was, all of them answered "the problem of the ethnic group." It is quite likely that this is because in this geographic region of Ecuador, the inhabitants are not familiar with their ethnic group, which is mostly montuvia or mestizo. In contrast, in the Sierra and Amazon regions, the inhabitants, a good proportion of whom are indigenous, who have historically grouped themselves into peoples and nationalities, even recognized in the country’s political constitution, have a much clearer understanding of the issue. To minimize errors in the responses to this variable, the interviewers were instructed to explain the issue in detail to the participants.

Table 5 shows the responses to the question about the ambiguity of the questionnaire. Since this was the second question, the interviewers had already previously explained to the participants who previously had doubts regarding ethnicity what it was about. Apparently, this explanation was sufficient for the majority of the participants to fully understand the questionnaire in this second question. This gave the necessary confidence to decide that the explanation that the interviewer would give to the participants about the question, with greater emphasis on the coastal region, would be sufficient to avoid errors.

Data management plans: All data will be securely stored in password-protected files, and access will be limited to the research team. Personal identifiers will not be collected. All data will be confidential; nowhere in the form will names or identification numbers of the users of the health system who agree to participate be requested. The survey will only begin when the participant has understood what the research is about and agrees to participate.

Data Analysis: The data will be handled as follows: the interviewers will collect the information using a Google Forms that will feed the database, then they will be downloaded into an Excel (Microsoft Office) matrix, to then be tabulated and coded. The coded matrix will be entered into the IBM SPSS statistical program. In this database, frequency distribution tables will be made and the access methods will be related to the waiting time.

When the data are collected, they will be published in the same journal that publishes our protocol. After that, that is, when the protocol is accepted for publication, the data collection process will begin. This is possible because the approval of the ethics committee has already been obtained. Data will be analyzed using descriptive statistics and inferential statistics.

Statistical Analysis: Statistical analysis will include the calculation of means and standard deviations for continuous variables and the calculation of frequencies and percentages for categorical variables. For the statistical analysis, it is intended to make a comparison of the average time of specialized medical care, both for an appointment with a specialist and for the procedure, to then relate it to the social, demographic, economic variables and especially with the way in which the user accessed the appointment. To do so, the normality of the Waiting Time variable will be tested with the Kolmogorov-Smirnov test. If the result is normal, parametric tests such as ANOVA (post hoc Tukey), T test and Pearson correlations will be used depending on the independent variable to be analyzed. If the result is not normal, Kruskal-Wallis, Mann-Whitney and Spearman will be used. The variable Form of Access will also be considered as dependent and will be analyzed with Pearson’s Chi square. This variable will allow the creation of a new, but dichotomous variable that will be called Standardized Access. For this, the response options to the variable Form of Access: Through a friend or family member and through a person who is not a friend or family member, will be considered as Not Standardized and, the options: through a referral from a doctor at a health center, through a specialist from this or another hospital and through an appointment at the hospital or by phone call, will be considered as Yes Standardized. With this variable, we intend to perform a logistic regression to obtain Odds Ratios that help us better analyze the relationship with the independent variables.

For all tests, a significance level of 0.05 will be used as a reference. The statistical program SPSS version 25 will be used.

## Ethics statement

This research project was reviewed and approved by the Ethics Committee for Research on Human Beings of ITSUP with the number 1718079732. The ethics committee is registered in the Office for Human Research Protections with the number RB00014260.

All users who participate in this research will have the possibility of refusing to participate. Those who agree to do so will be informed through an informed consent, about the objective of the study. This consent was approved by the Ethics Committee that approved the protocol and will be in the main part of the response registration form. Even if they have agreed to participate, people can stop doing so at any time after the survey has started. In that case, their registration will be deleted in the presence of the participant. Participants will not receive any compensation for their participation. In addition, they will be informed of an email address where they can request more information about the research if they require it.

The interviewers will be properly identified and will respect the decision of the participants. They will not ask to see the identification or ask about the medical problems that led users to request an appointment at the hospital, only about the specialty they attend. Since the diseases treated by each medical specialty are very varied, it is not possible to determine the specific medical condition of a person just by knowing what type of specialist they attend.

This protocol was written and conditioned at Protocols.io and can be reviewed at: https://dx.doi.org/10.17504/protocols.io.261ge5z7wg47/v1

## Data Availability

All relevant data are within the manuscript and its Supporting Information files. They are also available at https://dx.doi.org/10.17504/protocols.io.261ge5z7wg47/v1 [Protocols.io]

## Acknowledgments

We would like to thank the Research Center of the University Espíritu Santo, especially Dr. Fernando Espinoza and Patricia Valencia for their trust and support in the creation of this research protocol.

## Disclaimer

To receive approval from the ethics committee for this research, it was necessary to present a letter of interest from the institution where the research was to be conducted. The Ministry of Public Health did not provide the aforementioned letter, therefore, all data collection will be carried out outside the hospitals. As it is a public space, approval from this institution is not necessary.

## Discussion

Investigating waiting times and methods of access to specialized health services in public hospitals in Ecuador is crucial for several reasons. Waiting times for specialized medical consultations in Ecuador’s public hospitals can be significantly long, averaging 49 days, with some cases extending up to 180 days. These prolonged waiting periods can have serious implications for patient health outcomes and the overall efficiency of the healthcare system. Briones (2024) highlighted the existence of informal access methods to obtain specialized medical appointments in public hospitals in Manabí, Ecuador. Patients who accessed appointments through informal means (e.g., with help from family or friends working at the hospital) experienced shorter waiting times by up to 19 days compared to those who used formal procedures (8). While informal access has been mentioned anecdotally, its potential role in exacerbating inequities within the health system has yet to be systematically studied in Ecuador.

Unregulated access to the healthcare system may perpetuate and even increase social inequalities, especially in Latin America, where structural inequities deeply influence living conditions and health outcomes (13). Socioeconomic disparities are key drivers of health access inequalities, and frequent barriers include the ability to pay, geographic distance, availability of services, cultural or ethnic differences, communication challenges, and architectural obstacles (13). Unregulated access could potentially worsen these issues by allowing individuals with more resources or connections to access healthcare more quickly, leaving marginalized populations at a disadvantage. Despite various Latin American countries implementing health reforms since the 1990s aimed at strengthening healthcare systems and reducing inequalities in access and outcomes (14), significant disparities persist. This suggests that unregulated access, without adequate oversight, may not address health inequalities and could worsen them in the absence of targeted interventions.

Addressing these health inequalities effectively requires the implementation of both structuring and sectorial policies in Latin America (13). These policies must regulate access to healthcare while also addressing the social determinants of health, such as income, education, transportation, and living conditions. Furthermore, a human rights-based approach, which includes principles of accountability, meaningful participation, transparency, and non-discrimination, should be incorporated into health policies to promote equitable universal health coverage in the region (15)

Ecuador has a complex healthcare system, serving a heterogeneous population of almost 18 million people through both public and private hospitals (16). This protocol aims to provide valuable information on waiting times and access methods to specialized services, contributing to the planning of more equitable health policies. Investigating waiting times and methods of access could help identify areas for improvement in these essential healthcare services. Studying these factors is critical for addressing healthcare inequities, improving service delivery, and ultimately enhancing patient outcomes in a healthcare system that faces significant resource limitations. To do so, we are proposing a methodology that is flexible and can be adapted for use in other countries with similar health systems, potentially facilitating international comparisons in future studies.

After an exhaustive search of the scientific literature, we found no studies on how Ecuadorians access specialized health services. Although informal access to specialized services has been mentioned anecdotally, it has not been formally studied in Ecuador. This study seeks to determine whether this practice occurs and how it influences waiting times, providing crucial insights for health policy.

The *Instituto Nacional de Estadística y Censos* (INEC), through its national health and nutrition survey, collects data on waiting times, but only for the period after a patient arrives at the hospital for treatment (17). It does not capture the waiting time between obtaining an appointment and receiving care. This study will expand on this variable, providing new information for public health planning. Additionally, identifying which medical specialties have the longest waiting times could reveal gaps in the availability of specialists, informing the need for targeted interventions.

The choice to use surveys to collect data on waiting times mirrors the methodology employed by INEC, Ecuador’s official statistical authority. The questions and response options in this study are similar to those used by INEC but have been modified to include information that INEC does not cover, thereby addressing the specific research question. This approach ensures reliability in the data collected. The use of trained surveyors will also improve the accuracy of responses by ensuring participants fully understand the survey’s objectives. Conducting the survey digitally and providing a link to users was considered, but posed a significant risk of non-response or inaccurate data, especially among populations with limited internet access.

As described in the methodology, the survey is not designed to diagnose a condition or form a construct. Instead, it seeks to gather objective data on waiting times and access methods. Therefore, it was not necessary to validate the questionnaire as a tool for measuring relationships between multiple variables. Instead, we validated the clarity and comprehensibility of the questions and response options. As detailed in the methodology, the survey questions were well-understood across the regions surveyed. In areas like the coastal region of Ecuador, where participants do not commonly group themselves by ethnicity, the role of trained surveyors becomes especially important. These surveyors will explain any potentially unfamiliar terms, such as "ethnicity," to ensure consistent understanding across regions.

The questionnaire was designed using questions previously validated in national studies, but it has been adapted to include new variables specific to this research. A comprehension test was conducted in different regions of Ecuador to ensure that all respondents fully understood the questions. Waiting for patients outside hospitals will allow us to reach a broad range of participants, including those without internet access, the elderly who may have difficulties using technology, and individuals with disabilities. This approach will ensure equitable participation from a representative sample of the Ecuadorian healthcare system’s users. The survey will be conducted in multiple hospitals in the most densely populated provinces, such as Guayas, Pichincha, and Manabí. In other provinces, we plan to visit the single general or regional hospital available in each location.

It is planned to publish the results of the research in the same journal that agrees to publish the protocol.

## Limitations

We recognize that interviewers may face security risks due to the current violence in certain areas of Ecuador. To mitigate this risk, safety measures will be implemented, including the use of inconspicuous equipment and coordination with local authorities.

In addition, it will be a limitation not to be able to carry out the research in basic and highly specialized hospitals. However, these would not affect the fulfillment of the objective of the research because they were not contemplated from the beginning. Also, although the sample was calculated in a stratified manner by the number of patients who go to hospitals in a reference period, it was not possible to do so by social or economic strata. Since the Ministry of Health hospitals are free of charge, it will only be possible to measure the method of access and the waiting time in a specific population that does not represent the total Ecuadorian population.

## Dissemination plans

The protocol and the results of the research will be disseminated through publications in scientific journals. In the event of modifications during the protocol review process, these will be adapted to the questionnaire and the data collection procedure.

## Supporting Information

Since this is a cross-sectional study, the STROBE checklist will be used.

